# Variability in excess deaths across countries with different vulnerability during 2020-2023

**DOI:** 10.1101/2023.04.24.23289066

**Authors:** John P.A. Ioannidis, Francesco Zonta, Michael Levitt

## Abstract

Excess deaths provide total impact estimates of major crises, such as the COVID-19 pandemic. We evaluated excess death’s trajectories during 2020-2023 across countries with accurate death registration and population age structure data; and assessed relationships with economic indicators of vulnerability. Using the Human Mortality Database on 34 countries, excess deaths were calculated for 2020-2023 (to week 29, 2023) using 2017-2019 as reference, with weekly expected death calculations and adjustment for 5 age strata. Countries were divided into less and more vulnerable; the latter had per capita nominal GDP<$30,000, Gini>0.35 for income inequality and/or at least 2.5% of their population living in poverty. Excess deaths (as proportion of expected deaths, p%) were inversely correlated with per capita GDP (r=-0.60), correlated with proportion living in poverty (r=0.66) and modestly correlated with income inequality (r=0.45). Incidence rate ratio for deaths was 1.06 (95% confidence interval, 1.04-1.08) in the more versus less vulnerable countries. Excess deaths started deviating in the two groups after the first wave. Between-country heterogeneity diminished over time within each of the two groups. Less vulnerable countries had mean p%=-0.8% and 0.4% in 0-64 and >65 year-old strata while more vulnerable countries had mean p%=7.0% and 7.2%, respectively. Usually lower death rates were seen in children 0-14 years old during 2020-2023 versus pre-pandemic years. While the pandemic hit some countries earlier than others, country vulnerability dominated eventually the cumulative impact. Half of the analyzed countries witnessed no substantial excess deaths versus pre-pandemic levels, while the other half suffered major death tolls.

**Significance Statement:** Excess deaths during 2020-2023 reflect the direct and indirect effects of the COVID-19 pandemic and of the measures taken. Data from 34 countries with detailed death registration and allowing to adjust for changes in the age structure of the population over time show two groups, each with very different excess death outcomes. The 17 more vulnerable countries (those with per capita nominal GDP<$30,000, Gini>0.35 for income inequality and/or at least 2.5% of their population living in poverty) had very high excess deaths compared with 2017-2019, while the other 17 less vulnerable countries had deaths during 2020-2023 that were comparable to 2017-2019. Continuous monitoring of excess deaths helps understand how country vulnerability shapes long-term impacts.

## Introduction

Estimates of excess deaths are considered to offer an aggregate picture of the impact on overall mortality during the COVID-19 crisis.^1,2^ Excess deaths capture the composite of deaths due to SARS-CoV-2, indirect effects of the pandemic, and the effects of the measures taken during the crisis (both positive and negative). Three years after COVID-19 was proclaimed as a pandemic, one can have a mature picture of the evolution of the excess deaths in different countries over time. This is most reliable in countries that have adequately complete death registration data and information on the evolution of their population age structure, so that proper age-adjustments can be made.^3^ Performance of different countries may have varied markedly. Moreover, excess deaths’ trajectories over time have also shown diverse patterns across countries, as different countries witnessed more excess deaths during different periods.

Here, we evaluate excess death trajectories in the period between January 2020 and July 2023 in 34 countries with the most reliable data. We evaluate how and why these trajectories have diverged across different countries overall and within different age groups. We categorize these countries into groups of more and less vulnerable ones. Vulnerability is judged based on per capita gross domestic product (GDP), income inequality and proportion of population below poverty level, since the COVID-19 crisis disproportionately affected the poor and the disadvantaged in high-income countries.^4–6^ Examining excess mortality as a function of vulnerability can help gain insights in why some countries performed poorly and others did much better during the pandemic. It could also offer predictive insights for future pandemic challenges and lessons for pandemic preparedness.

## Results

### Cumulative excess deaths and vulnerability indicators

Table 1 shows the estimated cumulative excess deaths (p%, absolute number, and per million) during the entire pandemic period and the 3 vulnerability indicators for each of the 34 countries (scatterplot in Figure 1). The Pearson correlation coefficients of p% with GDP per capita, Gini coefficient, and percentage living in poverty were -0.60, 0.45, and 0.66. Corresponding Spearman rank correlation coefficients were -0.69, 0.29, and 0.62, respectively. The negative binomial regression showed that the incidence rate ratio for death was 1.06 (95% confidence interval 1.04-1.08, p<0.0001) for more vulnerable countries versus less vulnerable countries; each of the three economic variables was also significantly associated with death risk (p=0.004 for GDP per capita, p=<0.0001 for Gini, p<0.0001 for percentage living in poverty).

**Table 1.**
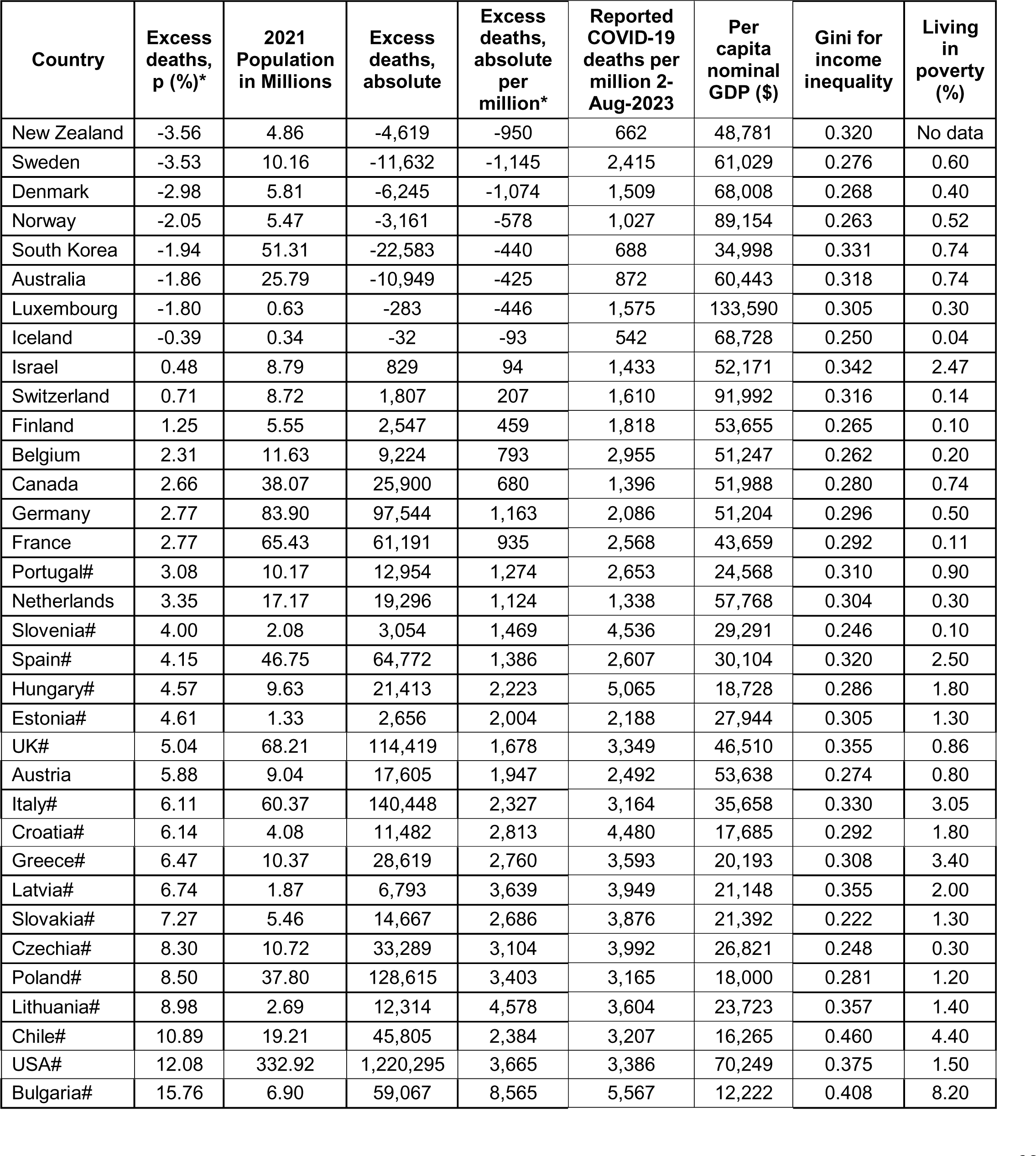
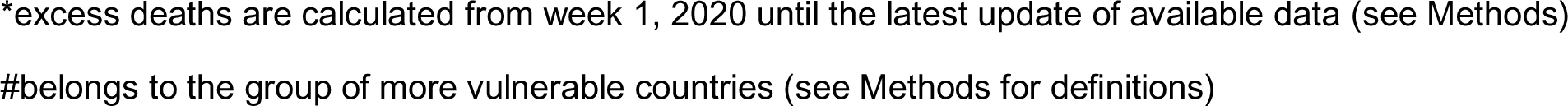
Excess deaths in 2020-2023 (using 2017-2019 average as reference) and vulnerability indicate.

| Country | Excess deaths, p (%)* | 2021 Population in Millions | Excess deaths, absolute | Excess deaths, absolute per million* | Reported COVID-19 deaths per million 2-Aug-2023 | Per capita nominal GDP (\$) | Gini for income inequality | Living in poverty (%) |
| --- | --- | --- | --- | --- | --- | --- | --- | --- |
| New Zealand | -3.56 | 4.86 | -4,619 | -950 | 662 | 48,781 | 0.320 | No data |
| Sweden | -3.53 | 10.16 | -11,632 | -1,145 | 2,415 | 61,029 | 0.276 | 0.60 |
| Denmark | -2.98 | 5.81 | -6,245 | -1,074 | 1,509 | 68,008 | 0.268 | 0.40 |
| Norway | -2.05 | 5.47 | -3,161 | -578 | 1,027 | 89,154 | 0.263 | 0.52 |
| South Korea | -1.94 | 51.31 | -22,583 | -440 | 688 | 34,998 | 0.331 | 0.74 |
| Australia | -1.86 | 25.79 | -10,949 | -425 | 872 | 60,443 | 0.318 | 0.74 |
| Luxembourg | -1.80 | 0.63 | -283 | -446 | 1,575 | 133,590 | 0.305 | 0.30 |
| Iceland | -0.39 | 0.34 | -32 | -93 | 542 | 68,728 | 0.250 | 0.04 |
| Israel | 0.48 | 8.79 | 829 | 94 | 1,433 | 52,171 | 0.342 | 2.47 |
| Switzerland | 0.71 | 8.72 | 1,807 | 207 | 1,610 | 91,992 | 0.316 | 0.14 |
| Finland | 1.25 | 5.55 | 2,547 | 459 | 1,818 | 53,655 | 0.265 | 0.10 |
| Belgium | 2.31 | 11.63 | 9,224 | 793 | 2,955 | 51,247 | 0.262 | 0.20 |
| Canada | 2.66 | 38.07 | 25,900 | 680 | 1,396 | 51,988 | 0.280 | 0.74 |
| Germany | 2.77 | 83.90 | 97,544 | 1,163 | 2,086 | 51,204 | 0.296 | 0.50 |
| France | 2.77 | 65.43 | 61,191 | 935 | 2,568 | 43,659 | 0.292 | 0.11 |
| Portugal# | 3.08 | 10.17 | 12,954 | 1,274 | 2,653 | 24,568 | 0.310 | 0.90 |
| Netherlands | 3.35 | 17.17 | 19,296 | 1,124 | 1,338 | 57,768 | 0.304 | 0.30 |
| Slovenia# | 4.00 | 2.08 | 3,054 | 1,469 | 4,536 | 29,291 | 0.246 | 0.10 |
| Spain# | 4.15 | 46.75 | 64,772 | 1,386 | 2,607 | 30,104 | 0.320 | 2.50 |
| Hungary# | 4.57 | 9.63 | 21,413 | 2,223 | 5,065 | 18,728 | 0.286 | 1.80 |
| Estonia# | 4.61 | 1.33 | 2,656 | 2,004 | 2,188 | 27,944 | 0.305 | 1.30 |
| UK# | 5.04 | 68.21 | 114,419 | 1,678 | 3,349 | 46,510 | 0.355 | 0.86 |
| Austria | 5.88 | 9.04 | 17,605 | 1,947 | 2,492 | 53,638 | 0.274 | 0.80 |
| Italy# | 6.11 | 60.37 | 140,448 | 2,327 | 3,164 | 35,658 | 0.330 | 3.05 |
| Croatia# | 6.14 | 4.08 | 11,482 | 2,813 | 4,480 | 17,685 | 0.292 | 1.80 |
| Greece# | 6.47 | 10.37 | 28,619 | 2,760 | 3,593 | 20,193 | 0.308 | 3.40 |
| Latvia# | 6.74 | 1.87 | 6,793 | 3,639 | 3,949 | 21,148 | 0.355 | 2.00 |
| Slovakia# | 7.27 | 5.46 | 14,667 | 2,686 | 3,876 | 21,392 | 0.222 | 1.30 |
| Czechia# | 8.30 | 10.72 | 33,289 | 3,104 | 3,992 | 26,821 | 0.248 | 0.30 |
| Poland# | 8.50 | 37.80 | 128,615 | 3,403 | 3,165 | 18,000 | 0.281 | 1.20 |
| Lithuania# | 8.98 | 2.69 | 12,314 | 4,578 | 3,604 | 23,723 | 0.357 | 1.40 |
| Chile# | 10.89 | 19.21 | 45,805 | 2,384 | 3,207 | 16,265 | 0.460 | 4.40 |
| USA# | 12.08 | 332.92 | 1,220,295 | 3,665 | 3,386 | 70,249 | 0.375 | 1.50 |
| Bulgaria# | 15.76 | 6.90 | 59,067 | 8,565 | 5,567 | 12,222 | 0.408 | 8.20 |

\*excess deaths are calculated from week 1, 2020 until the latest update of available data (see Methods)
#belongs to the group of more vulnerable countries (see Methods for definitions)

**Figure 1.**
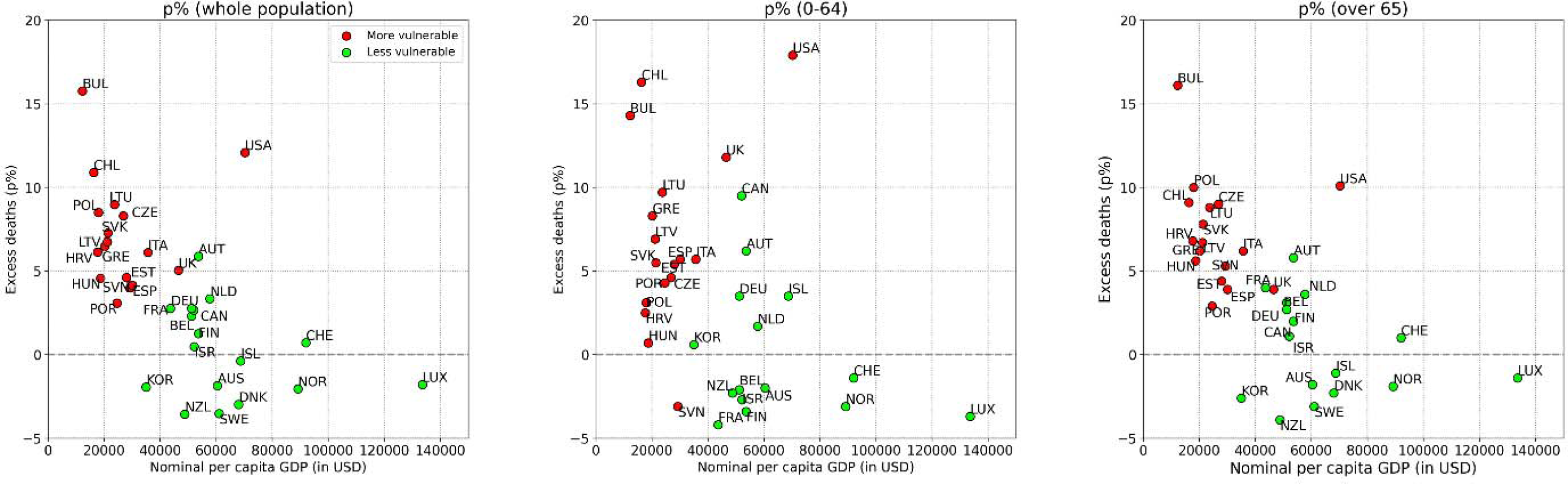
Scatterplot of excess deaths p% against the per capita GDP also showing countries with substantial income inequality and/or high proportion of population living in poverty. The left panel shows the data for the entire countries’ populations and the other two panels show the strata or non-elderly and elderly using a cut-off of 65 years.

Across the 34 countries there were 2,097,101 excess deaths, 58% were accounted by the USA alone (1,220,295). There were only 176,439 excess deaths in the 17 countries of the less vulnerable group (500 per million in the 2021 total population of 352,667,986, people) versus 1,920,662 in the 17 countries of the more vulnerable group (3,046 per million in the total population of 630,541,384 people).

The USA would have had 1.60 million fewer deaths if it had the performance of Sweden, 1.07 million fewer deaths if it had the performance of Finland, and 0.91 million fewer deaths if it had the performance of France.

### Cumulative excess deaths in the two vulnerability groups over time

Figure 2 shows country trajectories over time; means and standard deviations for p% for more and less vulnerable countries are in Table S1. Both groups had similar estimates of modest death deficit for the first 12 weeks of 2020 (versus the 2017-2019 average) and similar standard deviations. By mid-2020, both groups had reverted from this initial death deficit to a picture that resembled 2017-2019 for the mean p% (no sizeable excess or deficit), but there was large between-country heterogeneity already, especially in the more vulnerable group. Six countries had already reached excess deaths exceeding 8%.

**Figure 2.**
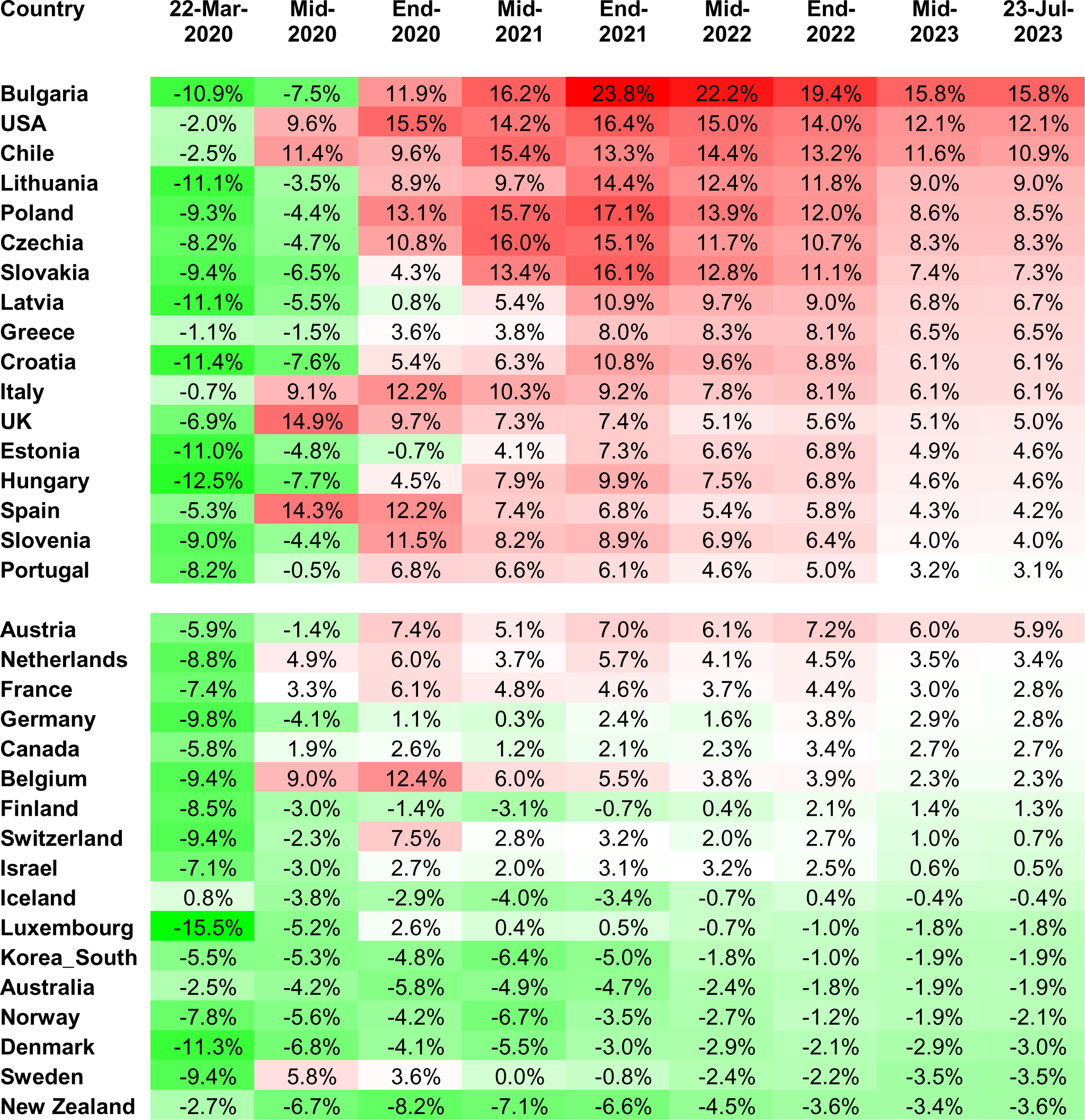
Estimated cumulative excess deaths at different timepoints during 2020-2023 in 34 countries grouped into more vulnerable (upper group) and less vulnerable (lower group). The color scale corresponds to excess deaths increasing from green to red.

After mid-2020, the less vulnerable group of countries maintained until July 2023 a very small mean of excess deaths (never exceeding 1.3%, 0.2% by the latest update). Their between-country heterogeneity also diminished during 2021-2023. The standard deviation at the latest update was half the standard deviation of end-2020 in that group. Countries that initially had prominent death deficits compared with 2017-2019 lost much or all of these death deficits by latest follow-up; while countries with substantial excess deaths in 2020 or early 2021 improved subsequently. As of the latest update in 2023, with the exception of Austria (p%=5.9%), all the less vulnerable countries had cumulative p% estimates in the narrow range -3.6% to 3.4%.

Conversely, among more vulnerable countries mean p% reached 8.2% by end-2020. Between-country heterogeneity diminished markedly compared with mid-2020, because although many of these countries were spared in the first wave, only Latvia and Estonia were still spared by end-2020 (standard deviation of p% fell from 8.2% to 4.6%). Mean p% continued to increase during 2021 reaching 9.9% by mid-2021 and 11.8% by end-2021, while between-country heterogeneity remained stable (between 4.4% and 4.8%). During 2022 and into 2023, mean p% decreased modestly over time and some further decrease was seen also in the standard deviation. As of the latest update in 2023, with the exception of Portugal (3.1%) all the more vulnerable countries had more cumulative excess deaths (4.0% to 15.8%) than 16 of the 17 less vulnerable countries.

There was no correlation between excess deaths p% and per capita GDP in 2020; this changed in 2021 and then remained steady between -0.55 and -0.62 (Table S2). Similarly, there was no correlation between excess deaths p% and the proportion of poverty until mid-2020 but this changed in the next year and then it remained steady between 0.5 and 0.66. Throughout 2020-2023 period, there was modest correlation (correlation coefficients mostly between 0.2 and 0.4) between excess deaths p% and income inequality.

### Sensitivity analyses using week-specific trends

In analyses using the week-specific trends method, as reported in STMF (Supplementary Table S3), the excess death estimates for most countries tended to be modestly higher than those obtained by considering as reference the average of 2017-2019 (Supplementary Figure S1). Nevertheless, the overall patterns (difference between less and more vulnerable, correlations with economic indicators, and evolution over time) remained largely unchanged. Countries in the two groups were separated by a cut-off of p%=9.1%, except for Netherlands (13.7%) and Austria (9.4%) having higher values in the less vulnerable group and Spain (8.1%), Lithuania (7.5%), and Italy (7%) having lower values in the more vulnerable group.

### Age groups

Across all 34 countries, 28% of total excess deaths were in the non-elderly (0-64 years) and 72% in the elderly (65 and over) strata (Table 2). For the non-elderly, at the latest update, excess deaths means were -0.8% and 7.0% in the less and more vulnerable groups, respectively. For the elderly, respective means were 0.4% and 7.2%. In the 17 less vulnerable countries, there were 13,472 and 162,967 excess deaths in the non-elderly and non-elderly strata, respectively; non-elderly excess deaths accounted for 7.6% of total excess deaths. In the 17 more vulnerable countries respective numbers were 577,281 and 1,343,381; non-elderly excess deaths accounted for 30% of total excess deaths.

**Table 2.**
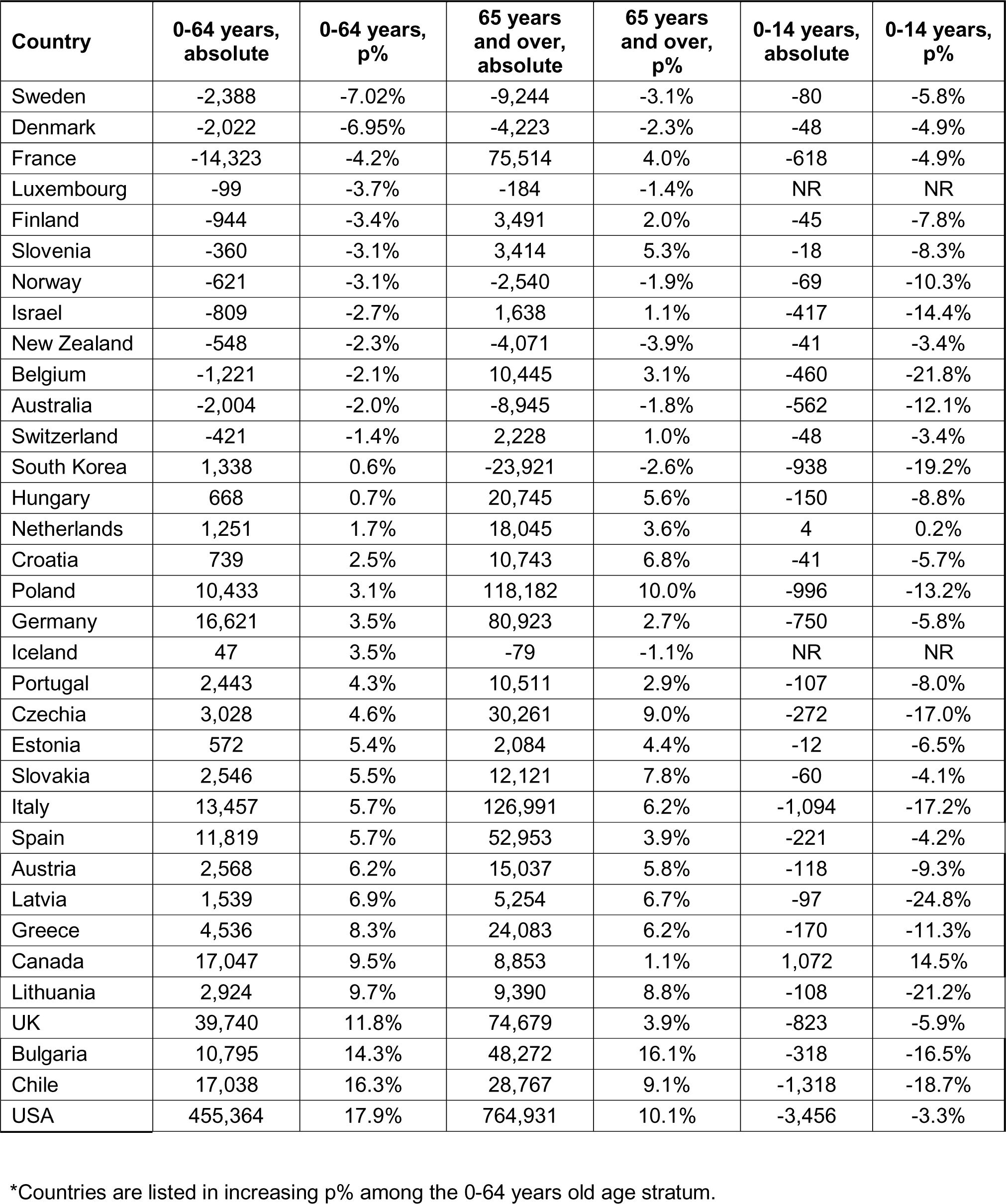
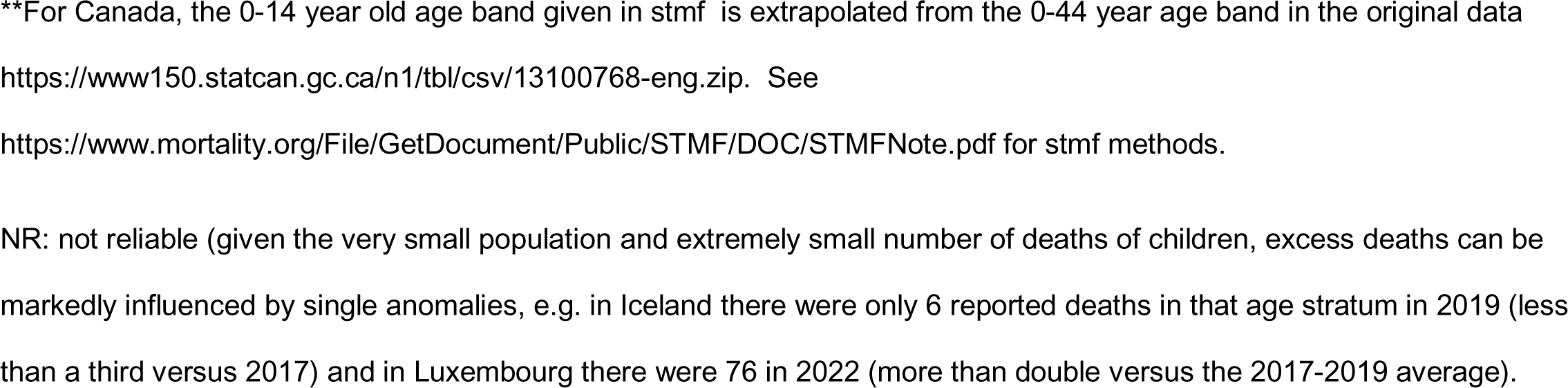
Excess deaths from January 2020 and up to latest update (week 29, 2023) per age group*.

| **Country** | **0-64 years,**<br>**absolute** | **0-64 years,**<br>**p%** | **65 years**<br>**and over,**<br>**absolute** | **65 years**<br>**and over,**<br>**p%** | **0-14 years,**<br>**absolute** | **0-14 years,**<br>**p%** |
| --- | --- | --- | --- | --- | --- | --- |
| Sweden | -2,388 | -7.02% | -9,244 | -3.1% | -80 | -5.8% |
| Denmark | -2,022 | -6.95% | -4,223 | -2.3% | -48 | -4.9% |
| France | -14,323 | -4.2% | 75,514 | 4.0% | -618 | -4.9% |
| Luxembourg | -99 | -3.7% | -184 | -1.4% | NR | NR |
| Finland | -944 | -3.4% | 3,491 | 2.0% | -45 | -7.8% |
| Slovenia | -360 | -3.1% | 3,414 | 5.3% | -18 | -8.3% |
| Norway | -621 | -3.1% | -2,540 | -1.9% | -69 | -10.3% |
| Israel | -809 | -2.7% | 1,638 | 1.1% | -417 | -14.4% |
| New Zealand | -548 | -2.3% | -4,071 | -3.9% | -41 | -3.4% |
| Belgium | -1,221 | -2.1% | 10,445 | 3.1% | -460 | -21.8% |
| Australia | -2,004 | -2.0% | -8,945 | -1.8% | -562 | -12.1% |
| Switzerland | -421 | -1.4% | 2,228 | 1.0% | -48 | -3.4% |
| South Korea | 1,338 | 0.6% | -23,921 | -2.6% | -938 | -19.2% |
| Hungary | 668 | 0.7% | 20,745 | 5.6% | -150 | -8.8% |
| Netherlands | 1,251 | 1.7% | 18,045 | 3.6% | 4 | 0.2% |
| Croatia | 739 | 2.5% | 10,743 | 6.8% | -41 | -5.7% |
| Poland | 10,433 | 3.1% | 118,182 | 10.0% | -996 | -13.2% |
| Germany | 16,621 | 3.5% | 80,923 | 2.7% | -750 | -5.8% |
| Iceland | 47 | 3.5% | -79 | -1.1% | NR | NR |
| Portugal | 2,443 | 4.3% | 10,511 | 2.9% | -107 | -8.0% |
| Czechia | 3,028 | 4.6% | 30,261 | 9.0% | -272 | -17.0% |
| Estonia | 572 | 5.4% | 2,084 | 4.4% | -12 | -6.5% |
| Slovakia | 2,546 | 5.5% | 12,121 | 7.8% | -60 | -4.1% |
| Italy | 13,457 | 5.7% | 126,991 | 6.2% | -1,094 | -17.2% |
| Spain | 11,819 | 5.7% | 52,953 | 3.9% | -221 | -4.2% |
| Austria | 2,568 | 6.2% | 15,037 | 5.8% | -118 | -9.3% |
| Latvia | 1,539 | 6.9% | 5,254 | 6.7% | -97 | -24.8% |
| Greece | 4,536 | 8.3% | 24,083 | 6.2% | -170 | -11.3% |
| Canada | 17,047 | 9.5% | 8,853 | 1.1% | 1,072 | 14.5% |
| Lithuania | 2,924 | 9.7% | 9,390 | 8.8% | -108 | -21.2% |
| UK | 39,740 | 11.8% | 74,679 | 3.9% | -823 | -5.9% |
| Bulgaria | 10,795 | 14.3% | 48,272 | 16.1% | -318 | -16.5% |
| Chile | 17,038 | 16.3% | 28,767 | 9.1% | -1,318 | -18.7% |
| USA | 455,364 | 17.9% | 764,931 | 10.1% | -3,456 | -3.3% |

\*Countries are listed in increasing p% among the 0-64 years old age stratum.
\*\*For Canada, the 0-14 year old age band given in stmf is extrapolated from the 0-44 year age band in the original data
https://www150.statcan.gc.ca/n1/tbl/csv/13100768-eng.zip. See
https://www.mortality.org/File/GetDocument/Public/STMF/DOC/STMFNote.pdf for stmf methods.
NR: not reliable (given the very small population and extremely small number of deaths of children, excess deaths can be markedly influenced by single anomalies, e.g. in Iceland there were only 6 reported deaths in that age stratum in 2019 (less than a third versus 2017) and in Luxembourg there were 76 in 2022 (more than double versus the 2017-2019 average)).

Most countries had slightly lower p% in the non-elderly and elderly strata (by 1.8%), with some notable exceptions (Table 2, Figure 1). Canada, UK, USA, and Chile had much higher p% in the non-elderly than the elderly (difference over 7.2%). USA had the highest excess deaths than any country among the non-elderly (p%=17.9%), while Canada had high excess deaths in the non-elderly (p%=9.5%), but no excess deaths in the elderly (p%=1.1%). Higher estimates in the non-elderly rather than elderly were manifest in these countries from the first pandemic months and persisted subsequently. Conversely, Slovenia, France, and Poland performed much better in the non-elderly than in the elderly (difference over 6.9%); France and Slovenia had death deficits in non-elderly.

For children (0-14 years), most countries had lower death rates during 2020-2023 versus 2017-2019. Numerically, the largest death deficits came actually from more vulnerable countries (USA (−3,456), Chile (−1,318), Italy (−1,094), Poland (−996)).

Supplementary Table S4 provides estimates of p%, absolute excess deaths and per million excess deaths in each country for each of the 2 non-elderly age strata (0-14, 15-64) and Supplementary Table S5 does the same for each of the 3 elderly strata (65-74, 75-84, 85+). Supplementary Table S6 provides standardized excess death rates based on the 2013 revision of the European Standard Population.

## Discussion

Analysis of data during 2020-2023 from 34 countries with reliable mortality data and information of population age structure shows that half of these countries had minimal excess deaths or even death deficits during 2020-2023 versus the 3 pre-pandemic years, while the other half had substantial excess deaths. Performance of different countries could be grouped based on vulnerability indicators pertaining to their wealth, income inequality and poverty. Excess deaths during 2020-2023 was strongly inversely correlated with per capita GDP, strongly correlated with proportion of population living in poverty and modestly correlated with income inequality. These correlational patterns were not seen during the first wave, but they became manifest and persistent subsequently.

Some less vulnerable countries transiently had notable excess deaths during 2020 and/or 2021. However, by mid-2023, all except Austria (and Netherlands in a week-specific trend sensitivity analysis, Figure S2) had no major excess deaths considering the entire 2020-2023 period. Perhaps transient peaks of excess deaths in these countries were to a large extent due to the demise of frail elderly individuals, e.g. in long-term care facilities, with limited life expectancy.^7^ For individuals with life expectancy less than 2 to 3 years, premature deaths in the early pandemic, contribute no excess deaths when the whole 2020-2023 period is considered.^7,8^ The group of less vulnerable countries have high income, no prominent income inequalities and no major proportion of their population living in poverty. All of them also have practically universal health coverage. Thus, they may have fewer excess deaths among disadvantaged people. These countries also had more means to mobilize sufficient healthcare and public health resources. The exact ingredients of their success cannot be deciphered from ecological, country-level data. Regardless, despite having many elderly, these countries managed to pass the crisis with no more deaths than during the recent pre-pandemic years, after adjusting for age structure.

Conversely, more vulnerable countries fared poorly during 2020-2023. Their excess death estimates have decreased modestly after peaking by end-2021. This may reflect a relative contribution of the same phenomenon of some early deaths among frail elderly people with limited life expectancy, especially in Slovenia, Slovakia, Poland, Spain and UK, where decreases in cumulative excess deaths were prominent over time. It may also reflect the more limited fatality of Omicron waves in 2022-2023,^9^ especially given almost ubiquitous prior infections and high rates of vaccination.^10^ However, cumulative excess death estimates remain high (and occasionally very high) in the more vulnerable countries. Some countries already reached high excess levels in early 2020, while others succumbed during the second wave. Again, the exact ingredients of their failure cannot be pinpointed in detail. However, the common denominator for these countries was their less robust economies and/or large share of poor, disadvantaged people. COVID-19 was a crisis of inequalities and many measures taken may even have fostered worsening inequalities.

The USA is a striking case, with extremely high cumulative excess death rates despite high per capita GDP. USA income inequality is high, many people live in poverty, and many lack health insurance coverage (among working-age people 15% in 2019 and 12% in 2022, among the entire population 10% in 2019 and 8% in 2022).^11^ Up to 27-fold differences in COVID-19 death rates in strata defined by race, gender, and educational achievement have been described in the USA.^6^ Area deprivation may be a risk factor for COVID-19 mortality especially among minorities.^12,13^ Moreover, many excess deaths apparently are due to sharply increasing deaths due to overdose^14^ and deaths due to suboptimal healthcare access during 2020-2023.^15^ Such non-infectious causes may largely explain the exceptionally high excess death p% estimate among USA non-elderly people.

Income inequalities have been strongly associated with COVID-19 deaths also in other vulnerable countries, e.g. in Spain^16,17^ and Chile.^18^ The pandemic widened all-cause pre-existing mortality gaps according to income even in some less vulnerable countries, e.g. Netherlands^19^ and Sweden.^20^ In a vicious circle, the pandemic caused more poverty worldwide.^21^

The gap between the more and less vulnerable group of countries was overall similar among the elderly and the non-elderly. However, Canada, UK, USA and Chile had very poor performance specifically among the non-elderly, while Slovenia, France, and Poland showed the opposite pattern. Population obesity rates are markedly different in these two groups of countries (28-36% versus 20-23%) and additional differences in background health status of non-elderly individuals may explain these patterns, e.g. overdose and poor healthcare access in the USA – as discussed above. USA, Chile, and UK were categorized here as more vulnerable countries, while Canada was categorized as less vulnerable. However, the classification is not absolute. Canada also has a modest proportion of people living in poverty (0.74%), and data from Ontario suggest that low income and low educational attainment increased risks of COVID-19 deaths.^22^ In British Columbia death rates from overdose more than doubled during the pandemic versus 2019.^23^

For children, most countries had fewer deaths during 2020-2023 compared with the 3 pre-pandemic years. This may be due to the exceptionally low infection fatality rate of SARS-CoV-2 among children,^24^ plus the almost complete disappearance of influenza for 2 years. More granular data should be examined to see if death deficits extend also to adolescents and very young adults.

Our study has some limitations. First, excess death estimates depend on modeling assumptions that may affect the absolute values.^25,26^ In a sensitivity analysis considering the SMTF week-specific trends with decreasing mortality rates over the years, the estimated excess deaths were higher than what we calculated using the 2017-2019 average as reference. However, even if mortality rates had decreased in many countries over time in the past, there is no guarantee that decreases should continue, especially among the increasingly elderly and frail populations of high-income countries.^27^ In essence, the analysis including linear trends based on past years assumes that a continued 2-5% decrease in death rate should continue to materialize every year in most of these high-income countries. When there is no increase in deaths, this is then counted as substantial excess mortality when calibrated against that optimistic extrapolation of continuous constant improvement. Furthermore, extrapolating the trend 4 years out from the end of the reference period may also be problematic for any trend analysis. Moreover, we have previously shown that different modeling assumptions do not affect the comparative performance of different countries:^27^ poor performers are consistently poor performers regardless of the exact modeling and this was largely borne out also in the current time-trend sensitivity analysis. Therefore, the group-level contrast between less and more vulnerable countries is probably robust to modeling choices. Of note, one should avoid making inferences about the relative ranking of countries in pairwise comparisons. The p% for each country are estimates and they carry large uncertainty, not only because of statistical chance error (which may be modeled by a Poisson or negative binomial process for expected deaths), but also additional uncertainty due to potential errors in the underlying data and multiplicity of options in the modeling assumptions. Instead, one should focus on the big picture of the relative performance of the two large groups of less and more vulnerable countries.

Second, some data may have imperfections, e.g. some missing deaths in the last few weeks of the covered period, or inaccuracies in inferred data for the population structure in each age bin and country over time. However, the data for the 34 chosen countries are likely to be the most reliable. Extrapolations to countries with less reliable death and age structure data are precarious.^28^ Socioeconomic factors may have had a major impact on mortality during 2020-2023 also in other countries, perhaps even to a larger extent than in the 34 countries analyzed here.^29–31^ Their impact might have been even larger for non-COVID-19 deaths.^29^ Conversely, most countries not analyzed here have much younger populations and few frail elderly in long-term care facilities than the 34 analyzed countries; thus deaths directly due to SARS-CoV-2 are expected to have been fewer.^24,32^

Third, we used three high-level economic indicators and these should be seen as surrogate of a mixture of many socioeconomic and other factors that operate at individual, household, work, community, and societal levels. Poor outcomes during the COVID-19 crisis are probably determined by a large web of inter-related factors.

Acknowledging these caveats, the country-level analyses offer a picture of how countries succeeded or failed in dealing with this major crisis. Half the countries went through the 3 crisis years without witnessing substantial excess deaths versus their 2017-2019 pre-pandemic levels, while others suffered major death tolls. Continuous monitoring of excess death patterns may be useful, given that the impact of the COVID-19 crisis and several measures taken may continue for several years.

## Materials and Methods

### Excess death calculations, eligible countries, and mortality data

We performed excess death calculations using methods similar to those we have used in our previously published work for 2020-2021.^3^ Briefly, we used the average of years 2017-2019 as baseline and included age-adjustment in the calculations, considering data in 5 age strata (0–14, 15–64, 65–75, 75–85, and >85 years old). For each age stratum, we obtained the average mortality, the number of deaths per million for the population of the specific age stratum and estimated the expected deaths during each week of the pandemic periods of interest, correcting for population size in the specific age stratum and summing expected deaths across population strata.

We used the Human Mortality Database (https://www.mortality.org),^33^ specifically the Short Term Mortality Fluctuations (STMF) file (https://www.mortality.org/Public/STMF/Outputs/stmf.csv) that includes weekly data. Only countries that have excellent death registration and include data with weekly deaths in the STMF during the period from 2017 until at least the second half of 2022 were considered, to avoid spurious changes in recorded deaths over time due to changes in death registration. STMF had data from 34 such eligible countries when we downloaded the database (August 2023). Data covered up to at least week 20 (at most up to week 29, 2023) for 32 countries, up to week 17 for Australia and up to week 8 for Canada.

We calculated weekly excess deaths and focused on cumulative excess deaths up to the end of each week starting from the first week of 2020. Calculations proceeded until the latest week with available death data in each country. The Human Mortality database uses ISO weeks that end on Sunday, e.g. the first week of 2020 ends on Sunday January 5, 2020. We expressed the excess death impact as percentage above expected deaths, p%.

All analyses use 2017-2019 as the reference period to calculate expected deaths. We have previously shown that relative country performance tends to be similar with shorter and longer windows for the reference period,^27^ even if exact absolute excess deaths may vary with different modeling options.^27^ There is also debate in the literature about if and how linear (or other) trends should be considered in the modeling, with some researchers arguing in favor of considering such trends.^34,35^ We have previously shown that once changes in the age pyramid are taken into account, extra consideration of trends may be unnecessary or even unwanted (e.g. if it extrapolates that future mortality trend patterns should continue in the future, even if they are saturated.^3,27^ Nevertheless, for comparison, we also provide excess death estimates for the whole 2020-2023 period using the week-specific trends method that is already implemented in STMF;^7,36^ all available consecutive years in the window 2000-2019 are considered for the reference period and week-specific linear trends are considered in this option.

### Classification of countries in vulnerability groups

We aimed to classify the 34 countries into two groups of 17 countries each using a parsimonious set of economic variables to separate a more vulnerable group from a less vulnerable group. We reasoned that per capita nominal GDP should be a most straightforward classifier to consider. However, we also reasoned that one should also allow for high vulnerability when, despite a high per capita nominal GPD, there is large income inequality and/or there are many people who are very poor and disadvantaged and thus would not be shielded by the services that can be offered otherwise to the majority of citizens. Indeed, it is well known that the Preston curve (the relationship between income and life expectancy)^37^ is almost flat after a certain income (i.e. further increases in income do not improve life expectancy) and that disadvantaged segments of the population can have much higher mortality, even in wealthy countries. Therefore, we defined a priori the more vulnerable countries’ group to include countries with per capita nominal GDP <$30,000, large income inequality (Gini>0.35) and/or sizeable percentage (≧2.5%) of population in poverty (defined as living with <$5.50 per day (2011 PPP values)). The group of less vulnerable countries had none of these vulnerability features. These three indicators may correlate with many other indicators of vulnerability that span the individual, household, work, community, and societal levels. They may also offer surrogacy for lack of health care resources for poor, disadvantaged segments of the population.

Information on these three indicators was obtained from World Bank for per capita nominal GDP (2021 estimates;^38^ OECD for Gini of income inequality (2020 or more recent estimates, “Income Distribution Database”. OECD.org. Measure: Gini (disposable income, post taxes and transfers), coupled with data from the World Bank for countries that are not OECD members;^39,40^ and Wikipedia (2019 for most recent data of proportion of population living in poverty).^41^

The magnitude of the differences across more and less vulnerable countries can be best appreciated if one estimates how many fewer deaths a more vulnerable country would have had if it had the same performance as a less vulnerable country. To obtain this number, the absolute excess deaths of the more vulnerable country are multiplied by the difference in p% between the compared countries. Illustratively, we present such absolute numbers of deaths that could have been saved for the USA, if it had the performance of Sweden, Finland, or France.

### Analyses

Cumulative excess deaths for each country were analyzed for each week starting with week 1, 2020 and until the most recent available data. For the few countries with missing data on the last few weeks in this interval, the last available value was carried forward. Main milestones were: week 12 (22-March 2020, at the start of the impact of the pandemic wave in Europe and the USA), mid-2020, end-2020, mid-2021, end-2021, mid-2022, end-2022, mid-2023, and latest available (up to at most week 29, 2023).

We examined Pearson and Spearman correlation coefficients for the correlation between p% at each main milestone and the three economic variables (GDP per capita, Gini coefficient, percentage of population living in poverty) across all 34 countries. We also performed a negative binomial regression (to allow for overdispersion) to examine the impact of vulnerable status and each of the 3 economic indicators, adjusting for exposure (population size), and expected deaths.

We performed similar analyses splitting excess death estimates in non-elderly (0-64 years, i.e. combining the 0-14 and 15-64 years old strata) and elderly people (65 years old and above, i.e. combining the 65-74, 75-84, and over 85 years old strata). We also examined children in particular (0-14 years). Estimates of p%, absolute and per million excess deaths for all granular age strata are provided in supplements so as to allow any type of standardized comparisons^42^ by interested researchers.

## Data Availability

All data are in the manuscript and in publicly available databases.

## Author Contributions

J.P.A.I. wrote the first draft of the paper. M.L. and J.P.A.I. ran the main analyses. All authors discussed the ideas, analyzed data, interpreted the data, edited the paper, and approved the final version.

## Classification

Biological Sciences; Medical Sciences.

## Supporting Information

**Fig. S1.**
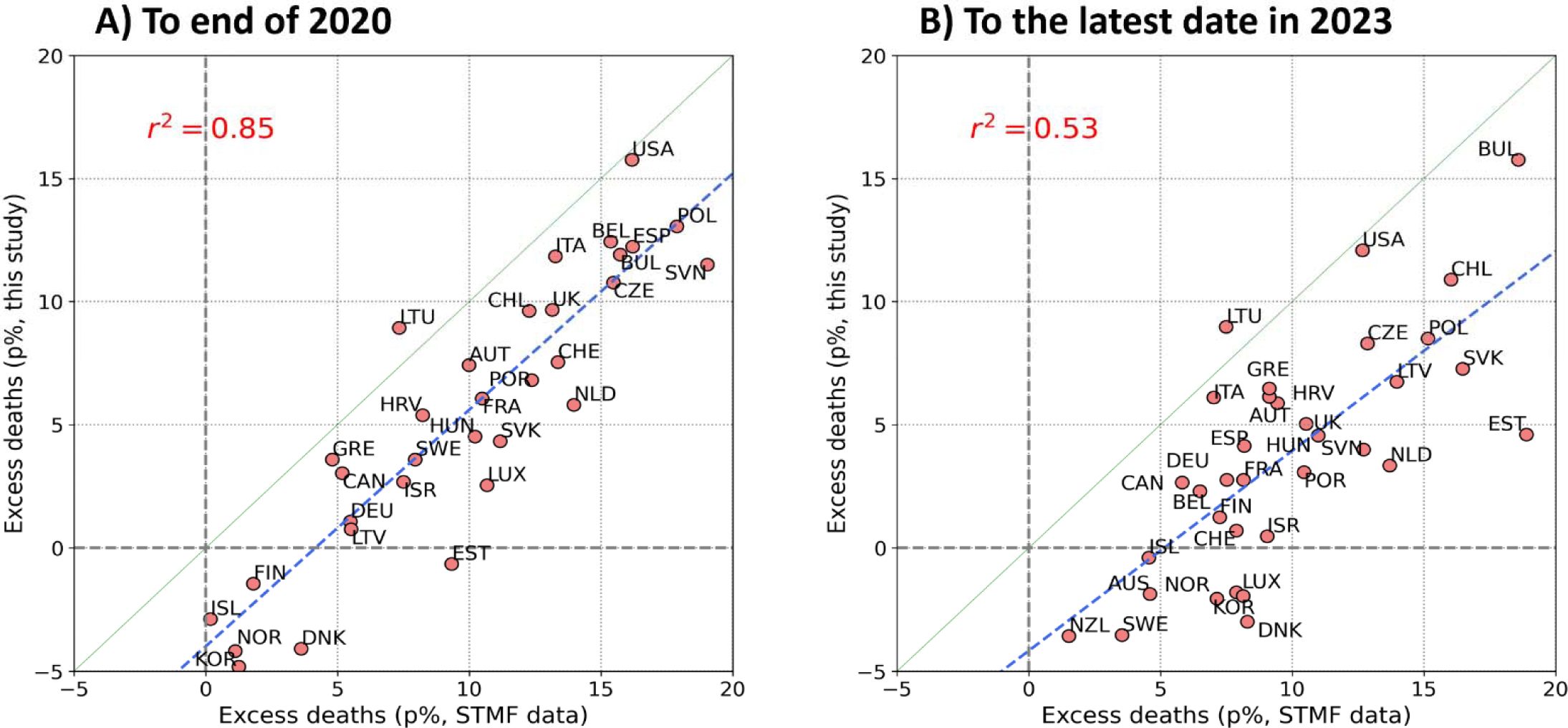
Scatterplot of excess deaths p%. A. until the end of 2020 and B. over the entire 2020-2023 period (to latest a ailable date in 2023) using the average of 2017-2019 as reference versus using the week-specific trend method with a linear trend^7^ and considering the 2000-2019 period (or longest span of consecutive years if start year with available data is later than 2000) as reference (estimates from STMF).

**Table S1.** Mean and standard deviation of excess deaths p% (% of expected deaths) at different time points for more and less vulnerable countries

| **Time point** | **Excess deaths in less vulnerable countries, mean p% (standard deviation)** | **Excess deaths in more vulnerable countries, mean p% (standard deviation)** |
| --- | --- | --- |
| March 2020 (week 12) | -7.4 (3.7) | -7.7 (3.9) |
| Mid-2020 (week 26) | -1.6 (4.7) | 0 (8.1) |
| End-2020 (week 52) | 1.2 (5.6) | 8.2 (4.5) |
| Mid-2021 (week 78) | -0.7 (4.4) | 9.8 (4.3) |
| End-2021 (week 104) | 0.3 (4.2) | 11.8 (4.7) |
| Mid-2022 (week 130) | 0.5 (3) | 10.2 (4.5) |
| End-2022 (week 156) | 1.2 (3) | 9.5 (3.7) |
| Mid-2023 (week 182) | 0.3 (2.8) | 7.3 (3.3) |
| July 2023 (week 185) | 0.2 (2.7) | 7.2 (3.3) |

**Table S2.**
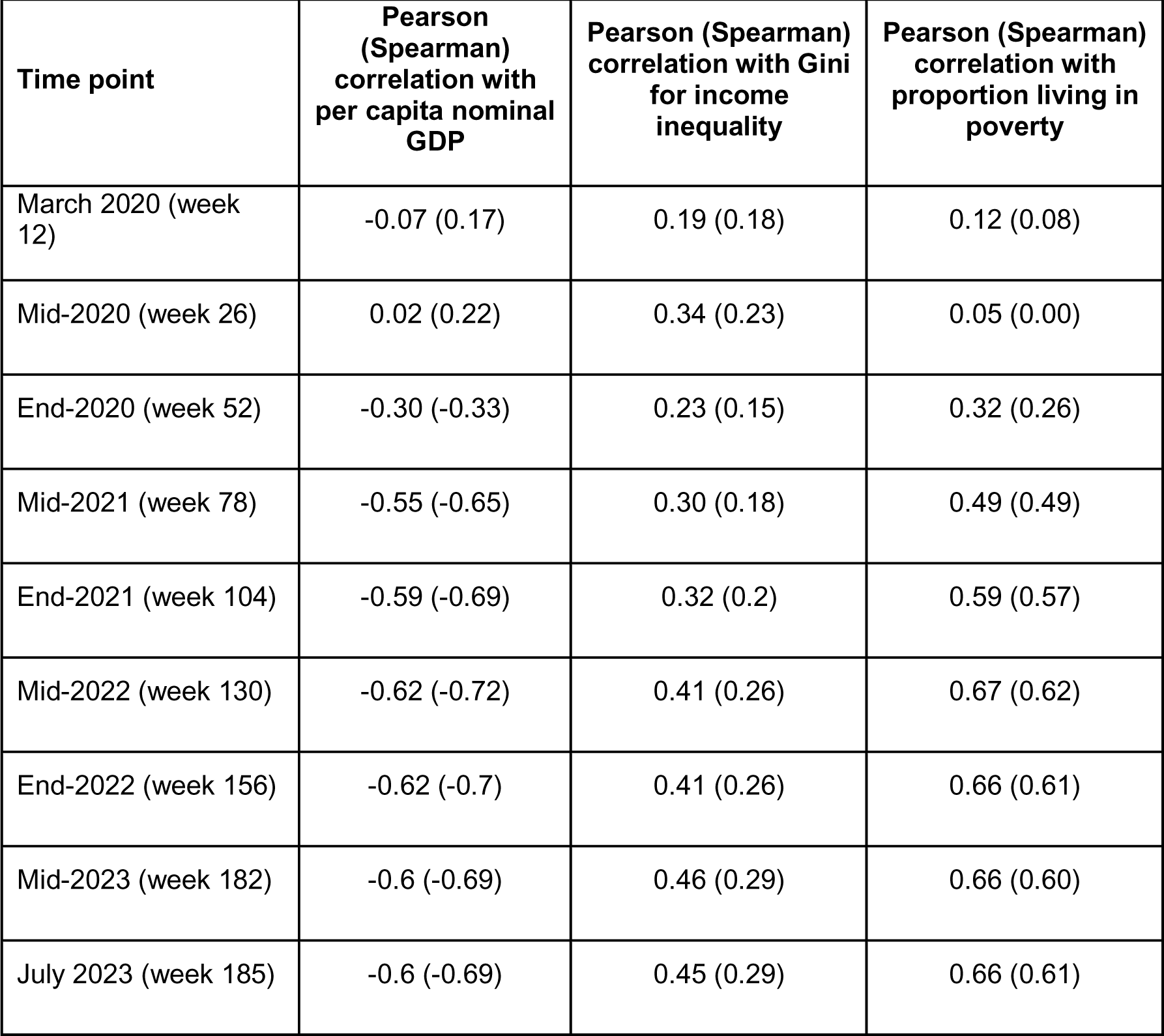
Correlations between excess death estimates p% and vulnerability indicators at different time points

| **Time point** | **Pearson (Spearman) correlation with per capita nominal GDP** | **Pearson (Spearman) correlation with Gini for income inequality** | **Pearson (Spearman) correlation with proportion living in poverty** |
| --- | --- | --- | --- |
| March 2020 (week 12) | -0.07 (0.17) | 0.19 (0.18) | 0.12 (0.08) |
| Mid-2020 (week 26) | 0.02 (0.22) | 0.34 (0.23) | 0.05 (0.00) |
| End-2020 (week 52) | -0.30 (-0.33) | 0.23 (0.15) | 0.32 (0.26) |
| Mid-2021 (week 78) | -0.55 (-0.65) | 0.30 (0.18) | 0.49 (0.49) |
| End-2021 (week 104) | -0.59 (-0.69) | 0.32 (0.2) | 0.59 (0.57) |
| Mid-2022 (week 130) | -0.62 (-0.72) | 0.41 (0.26) | 0.67 (0.62) |
| End-2022 (week 156) | -0.62 (-0.7) | 0.41 (0.26) | 0.66 (0.61) |
| Mid-2023 (week 182) | -0.6 (-0.69) | 0.46 (0.29) | 0.66 (0.60) |
| July 2023 (week 185) | -0.6 (-0.69) | 0.45 (0.29) | 0.66 (0.61) |

**Table S3.** Estimates of p% from 1/2020 to the end of each calendar year and to the latest available 2023 data according to the week-specific trend analyses provided in the SMTF (ref. 7 and ref. 11)

| **Country** | **End-2020** | **End-2021** | **End-2022** | **Latest-2023** | **Reference period start year** |
| --- | --- | --- | --- | --- | --- |
| Australia | -2.0% | 0.2% | 4.4% | 4.6% | 2015 |
| Austria | 10.0% | 9.7% | 10.3% | 9.4% | 2000 |
| Belgium | 15.4% | 8.8% | 7.8% | 6.5% | 2000 |
| Bulgaria | 15.7% | 27.7% | 22.4% | 18.6% | 2000 |
| Canada | 5.2% | 4.9% | 6.6% | 5.8% | 2010 |
| Chile | 12.3% | 16.9% | 17.7% | 16.0% | 2016 |
| Croatia | 8.2% | 13.3% | 11.6% | 9.1% | 2001 |
| Czechia | 15.5% | 19.7% | 15.2% | 12.8% | 2005 |
| Denmark | 3.6% | 6.0% | 8.3% | 8.3% | 2007 |
| Estonia | 9.3% | 19.0% | 20.0% | 18.9% | 2000 |
| Finland | 1.8% | 3.8% | 7.6% | 7.2% | 2000 |
| France | 10.5% | 9.4% | 9.6% | 8.1% | 2000 |
| Germany | 5.5% | 5.2% | 8.1% | 7.5% | 2000 |
| Greece | 4.8% | 9.7% | 10.2% | 9.1% | 2015 |
| Hungary | 10.2% | 16.0% | 13.1% | 11.0% | 2000 |
| Iceland | 0.2% | 0.4% | 4.8% | 4.5% | 2000 |
| Israel | 7.5% | 9.3% | 10.2% | 9.0% | 2000 |
| Italy | 13.3% | 10.1% | 9.0% | 7.0% | 2011 |
| South Korea | 1.3% | 2.6% | 8.5% | 8.1% | 2010 |
| Latvia | 5.5% | 16.6% | 15.7% | 14.0% | 2000 |
| Lithuania | 7.3% | 12.5% | 10.1% | 7.5% | 2000 |
| Luxembourg | 10.7% | 9.2% | 8.3% | 7.9% | 2000 |
| Netherlands | 14.0% | 14.4% | 14.2% | 13.7% | 2000 |
| New Zealand | -4.9% | -2.6% | 1.2% | 1.5% | 2010 |
| Norway | 1.1% | 3.3% | 7.2% | 7.1% | 2000 |
| Poland | 17.9% | 22.7% | 18.3% | 15.1% | 2000 |
| Portugal | 12.4% | 12.9% | 12.9% | 10.4% | 2000 |
| Slovakia | 11.2% | 24.0% | 19.9% | 16.5% | 2000 |
| Slovenia | 19.0% | 16.8% | 14.7% | 12.7% | 2000 |
| Spain | 16.2% | 10.8% | 9.9% | 8.2% | 2000 |
| Sweden | 7.9% | 4.5% | 4.3% | 3.5% | 2000 |
| Switzerland | 13.4% | 9.3% | 9.5% | 7.9% | 2000 |
| UK | 13.1% | 11.3% | 10.2% | 10.5% | 2015 |
| USA | 16.2% | 16.7% | 14.3% | 12.7% | 2015 |

**Table S4.** p%, absolute and per million excess deaths in non-elderly age strata (0-14, 15-64)

| Country | Excess deaths for 0-14 |  |  | Excess deaths for 15-64 |  |  |
| --- | --- | --- | --- | --- | --- | --- |
|  | p (%) | absolute | per million | p (%) | absolute | per million |
| Australia | -12.1 | -562 | -118.4 | -1.53 | -1442 | -86.6 |
| Austria | -9.3 | -118 | -92.5 | 6.6 | 2,687 | 454.7 |
| Belgium | -21.8 | -460 | -237.4 | -1.4 | -760 | -103.1 |
| Bulgaria | -16.5 | -318 | -318.3 | 15.1 | 11,114 | 2,509.9 |
| Canada | 14.5 | 1,072 | 178.1 | 9.3 | 15,975 | 637.0 |
| Chile | -18.7 | -1,318 | -379.6 | 18.8 | 18,357 | 1,507.1 |
| Croatia | -5.7 | -41 | -71.3 | 2.7 | 780 | 298.9 |
| Czechia | -17.0 | -272 | -161.4 | 5.2 | 3,300 | 491.3 |
| Denmark | -4.9 | -48 | -51.4 | -7.0 | -1,973 | -531.5 |
| Estonia | -6.5 | -12 | -57.2 | 5.6 | 585 | 696.1 |
| Finland | -7.8 | -45 | -52.7 | -3.3 | -898 | -262.7 |
| France | -4.9 | -618 | -53.9 | -4.2 | -13,705 | -340.0 |
| Germany | -5.8 | -750 | -65.6 | 3.7 | 17,371 | 324.5 |
| Greece | -11.3 | -170 | -112.9 | 8.9 | 4,707 | 693.9 |
| Hungary | -8.8 | -150 | -105.7 | 0.9 | 818 | 128.5 |
| Iceland | NR | NR | NR | 2.0 | 26 | 108.0 |
| Israel | -14.4 | -417 | -161.2 | -1.5 | -391 | -71.4 |
| Italy | -17.2 | -1,094 | -142.5 | 6.3 | 14,551 | 384.5 |
| South Korea | -19.2 | -938 | -147.0 | 1.0 | 2,276 | 61.8 |
| Latvia | -24.8 | -97 | -320.1 | 7.5 | 1,637 | 1,358.1 |
| Lithuania | -21.2 | -108 | -255.8 | 10.2 | 3,033 | 1,669.9 |
| Luxembourg | NR | 52 | NR | -5.8 | -152 | -348.0 |
| Netherlands | 0.2 | 4 | 1.6 | 1.7 | 1,247 | 110.4 |
| New Zealand | -3.4 | -41 | -42.7 | -2.2 | -507 | -153.1 |
| Norway | -10.3 | -69 | -74.8 | -2.8 | -552 | -157.8 |
| Poland | -13.2 | -996 | -169.1 | 3.5 | 11,430 | 449.8 |
| Portugal | -8.0 | -107 | -77.2 | 4.6 | 2,550 | 386.0 |
| Slovakia | -4.1 | -60 | -70.1 | 5.8 | 2,606 | 709.4 |
| Slovenia | -8.3 | -18 | -59.7 | -3.0 | -341 | -252.9 |
| Spain | -4.2 | -221 | -32.4 | 6.0 | 12,041 | 385.7 |
| Sweden | -5.8 | -80 | -43.7 | -7.1 | -2,308 | -358.4 |
| Switzerland | -3.4 | -48 | -37.6 | -1.3 | -372 | -65.2 |
| UK | -5.9 | -823 | -69.4 | 12.5 | 40,564 | 957.8 |
| United States | -3.3 | -3,456 | -56.7 | 18.8 | 458,821 | 2,132.2 |

NR: not reliable due to small numbers (see discussion in Table 2)

**Table S5.** p%, absolute and per million excess deaths in elderly age strata (65-74, 75-84, 85+)

| Country | Excess deaths for 65-74 |  |  | Excess deaths for 75-84 |  |  | Excess deaths for 85 Plus |  |  |
| --- | --- | --- | --- | --- | --- | --- | --- | --- | --- |
|  | p (%) | absolute | per million | p (%) | absolute | per million | p (%) | absolute | per million |
| Australia | -2.3 | -2,181 | -921.8 | -3.48 | -5564 | -4302.6 | -0.5 | -1198 | -2308.7 |
| Austria | 2.7 | 1,285 | 1,498.7 | 10.41 | 8933 | 14304.2 | 3.88 | 4817 | 21263.1 |
| Belgium | 4.0 | 2,560 | 2,163.5 | 1.23 | 1305 | 1864.4 | 3.88 | 6579 | 19657.8 |
| Bulgaria | 24.2 | 20,537 | 23,255.3 | 13.59 | 16084 | 33660.4 | 12.11 | 11650 | 79215.5 |
| Canada | 1.4 | 2,451 | 620.7 | 0.33 | 838 | 411.9 | 1.55 | 5562 | 6496.1 |
| Chile | 10.0 | 8,243 | 6,315.0 | 9.79 | 10790 | 15698.6 | 7.88 | 9732 | 45783.1 |
| Croatia | 11.5 | 4,232 | 8,808.3 | 5.74 | 3420 | 12004.6 | 5.1 | 3090 | 32613.9 |
| Czechia | 9.8 | 9,043 | 7,027.6 | 8.16 | 10260 | 15752.2 | 9.37 | 10957 | 53482.7 |
| Denmark | -0.9 | -324 | -505.9 | -3.92 | -2687 | -6718.1 | -1.65 | -1212 | -9591.6 |
| Estonia | 4.4 | 498 | 3,489.0 | 4.62 | 738 | 8178.1 | 4.28 | 847 | 23913.5 |
| Finland | 4.2 | 1,528 | 2,147.0 | -0.89 | -527 | -1392.7 | 3.1 | 2490 | 16249.7 |
| France | 4.9 | 16,667 | 2,278.1 | -1.72 | -8785 | -2180.9 | 6.61 | 67633 | 30138.9 |
| Germany | 4.4 | 23,916 | 2,745.2 | 7.04 | 73654 | 10468.3 | -1.14 | -16647 | -6793.7 |
| Greece | 9.7 | 5,957 | 5,041.8 | 2.8 | 3549 | 4254.51 | 7.3 | 14575 | 38147.9 |
| Hungary | 10.7 | 11,881 | 10,361.3 | 4.67 | 6460 | 10468.0 | 1.99 | 2403 | 11987.5 |
| Iceland | 2.2 | 28 | 929.4 | -6.54 | -158 | -10033 | 1.51 | 50 | 7687.6 |
| Israel | 3.8 | 1,144 | 1,729.1 | -1.92 | -903 | -2879.2 | 2.07 | 1398 | 10507.5 |
| Italy | 7.0 | 21,251 | 3,099.1 | 6.34 | 41637 | 8576.3 | 5.83 | 64102 | 29131.4 |
| South Korea | -5.7 | -10,814 | -2,332.4 | -2.54 | -9201 | -3390.3 | -1.03 | -3906 | -4975.4 |
| Latvia | 7.8 | 1,528 | 7,542.4 | 8.13 | 2371 | 16624.1 | 4.56 | 1353 | 28503.2 |
| Lithuania | 10.9 | 2,734 | 9,667.3 | 9.97 | 3776 | 18980.8 | 6.55 | 2878 | 38447.1 |
| Luxembourg | -7.7 | -209 | -4,152.1 | -3.93 | -165 | -5692.4 | 3.16 | 191 | 15560.1 |
| Netherlands | 3.7 | 3,674 | 1,882.3 | 3.9 | 6683 | 6154.1 | 3.33 | 7687 | 19776.5 |
| New Zealand | -4.1 | -904 | -1,972.6 | -5.19 | -1901 | -7811.0 | -2.69 | -1265 | -14298 |
| Norway | -2.4 | -614 | -1,142.3 | -5.21 | -2371 | -7920.3 | 0.71 | 445 | 3793.2 |
| Poland | 13.9 | 49,720 | 11,486.6 | 8.27 | 30686 | 16079.7 | 8.41 | 37775 | 45940.1 |
| Portugal | 3.6 | 2,157 | 1,827.9 | -0.78 | -958 | -1214.9 | 5.17 | 9311 | 26820.1 |
| Slovakia | 12.8 | 6,131 | 10,712.7 | 8.22 | 4702 | 17840.5 | 2.55 | 1287 | 15261.6 |
| Slovenia | 4.7 | 648 | 2,738.0 | 4.74 | 1017 | 7377.4 | 5.97 | 1749 | 31830.8 |
| Spain | 6.7 | 13,998 | 2,988.8 | -0.26 | -1052 | -343.4 | 5.41 | 40006 | 25406 |
| Sweden | -3.4 | -1,673 | -1,529.7 | -3.84 | -3969 | -5508.6 | -2.52 | -3600 | -13674 |
| Switzerland | -1.2 | -410 | -492.5 | -0.58 | -397 | -717.1 | 2.5 | 3037 | 13120.5 |
| UK | 7.0 | 25,601 | 3,812.1 | 2.56 | 17318 | 4187.7 | 3.57 | 31758 | 19011.8 |
| United States | 12.9 | 263,333 | 8,089.1 | 10.23 | 258053 | 16171 | 8.16 | 243543 | 40198.7 |

**Table S6.** Standardized excess death rates for 2020-2023 using the average of 2017-2019 as reference and using the 2013 European Standard Population

| **Country** | **Standardized excess death rate (per 100,000 population)** |
| --- | --- |
| Australia | -41 |
| Austria | 174 |
| Belgium | 51 |
| Bulgaria | 574 |
| Canada | 63 |
| Chile | 308 |
| Croatia | 178 |
| Czechia | 265 |
| Denmark | -103 |
| Estonia | 157 |
| Finland | 14 |
| France | 38 |
| Germany | 71 |
| Greece | 166 |
| Hungary | 105 |
| Iceland | -39 |
| Israel | 0 |
| Italy | 151 |
| South Korea | -33 |
| Latvia | 262 |
| Lithuania | 323 |
| Luxembourg | -21 |
| Netherlands | 97 |
| New Zealand | -97 |
| Norway | -53 |
| Poland | 246 |
| Portugal | 83 |
| Slovakia | 199 |
| Slovenia | 110 |
| Spain | 86 |
| Sweden | -94 |
| Switzerland | 23 |
| UK | 135 |

The 2013 European Standard Population has 16%, 64.5%, 10.5%, 6.5%, and 2.5% of the population in the 0-14, 15-64, 65-74, 75-84, and 85 and above years strata, respectively.

## References

1. Kiang MV, Irizarry RA, Buckee CO, Balsari S. Every body counts: measuring mortality from the COVID-19 pandemic. Ann Intern Med. 2020;173(12):1004–1007.

2. Msemburi W, Karlinsky A, Knutson V, Aleshin-Guendel S, Chatterji S, Wakefield J. The WHO estimates of excess mortality associated with the COVID-19 pandemic. Nature. 2023;613(7942):130–137.

3. Levitt M, Zonta F, Ioannidis JPA. Comparison of pandemic excess mortality in 2020-2021 across different empirical calculations. Envir Res. 2022;213:113754.

4. Shiels MS, Haque AT, Haozous EA, Albert PS, Almeida JS, García-Closas M, Nápoles AM, Pérez-Stable EJ, Freedman ND, Berrington de González A. Racial and ethnic disparities in excess deaths during the COVID-19 pandemic, March to December 2020. Ann Intern Med. 2021;174(12):1693–1699.

5. Commodore-Mensah Y, Cooper LA. Reversing the tide of racial and ethnic disparities in excess deaths during the COVID-19 pandemic. Ann Intern Med. 2021;174(12):1755–1756.

6. Pathak EB, Menard JM, Garcia RB, Salemi JL. Joint effects of socioeconomic position, race/ethnicity, and gender on COVID-19 mortality among working-age adults in the United States. Int J Environ Res Public Health. 2022;19(9):5479.

7. Ballin M, Ioannidis JP, Bergman J, Kivipelto M, Nordström A, Nordström P. Time-varying risk of death after SARS-CoV-2 infection in Swedish long-term care facility residents: a matched cohort study. BMJ Open. 2022;12(11):e066258.

8. Espuny Pujol F, Hancock R, Morciano M. Trends in survival of older care home residents in England: a 10-year multi-cohort study. Soc Sci Med 2021;282:113883.

9. Friis NU, Martin-Bertelsen T, Pedersen RK, Nielsen J, Krause TG, Andreasen V, Vestergaard LS. COVID-19 mortality attenuated during widespread Omicron transmission, Denmark, 2020 to 2022. Euro Surveill. 2023;28(3):2200547.

10. Pilz S, Ioannidis JPA. Does natural and hybrid immunity obviate the need for frequent vaccine boosters against SARS-CoV-2 in the endemic phase? Eur J Clin Invest. 2023;53(2):e13906.

11. Cohen RA, Cha AE. Health Insurance Coverage: Early Release of Estimates From the National Health Interview Survey, 2022. In: https://www.cdc.gov/nchs/data/nhis/earlyrelease/insur202305_1.pdf, last accessed August 8, 2023.

12. Magesh S, John D, Li WT, Li Y, Mattingly-App A, Jain S, Chang EY, Ongkeko WM. Disparities in COVID-19 outcomes by race, ethnicity, and socioeconomic status: A systematic-review and meta-analysis. JAMA Netw Open. 2021;4(11):e2134147.

13. Matthay EC, Duchowny KA, Riley AR, Thomas MD, Chen YH, Bibbins-Domingo K, Glymour MM. Occupation and educational attainment characteristics associated with COVID-19 mortality by race and ethnicity in California. JAMA Netw Open. 2022;5(4):e228406.

14. https://www.cdc.gov/nchs/pressroom/nchs_press_releases/2022/202205.htm, last accessed March 26, 2023.

15. Roth GA, Vaduganathan M, Mensah GA. Impact of the COVID-19 pandemic on cardiovascular health in 2020: JACC State-of-the-Art Review. J Am Coll Cardiol. 2022;80(6):631–640.

16. Politi J, Martín-Sánchez M, Mercuriali L, Borras-Bermejo B, Lopez-Contreras J, Vilella A, Villar J; COVID-19 Surveillance Working Group of Barcelona; Orcau A, de Olalla PG, Rius C. Epidemiological characteristics and outcomes of COVID-19 cases: mortality inequalities by socio-economic status, Barcelona, Spain, 24 February to 4 May 2020. Euro Surveill. 2021;26(20):2001138.

17. Fernández-Martínez NF, Ruiz-Montero R, Gómez-Barroso D, Rodríguez-Torronteras A, Lorusso N, Salcedo-Leal I, Sordo L. Socioeconomic differences in COVID-19 infection, hospitalisation and mortality in urban areas in a region in the South of Europe. BMC Public Health. 2022;22(1):2316.

18. Mena G, Aburto JM. Unequal impact of the COVID-19 pandemic in 2020 on life expectancy across urban areas in Chile: a cross-sectional demographic study. BMJ Open. 2022;12(8):e059201.

19. Wouterse B, Geisler J, Bär M, van Doorslaer E. Has COVID-19 increased inequality in mortality by income in the Netherlands? J Epidemiol Community Health. 2023;77(4):244–251.

20. Bell M, Hergens MP, Fors S, Tynelius P, de Leon AP, Lager A. Individual and neighborhood risk factors of hospital admission and death during the COVID-19 pandemic: a population-based cohort study. BMC Med. 2023;21(1):1.

21. Decerf B, Ferreira FHG, Mahler DG, Sterck O. Lives and livelihoods: Estimates of the global mortality and poverty effects of the Covid-19 pandemic. World Dev. 2021;146:105561.

22. Wang L, Calzavara A, Baral S, Smylie J, Chan AK, Sander B, Austin PC, Kwong JC, Mishra S. Differential patterns by area-level social determinants of health in coronavirus disease 2019 (COVID-19)-related mortality and non-COVID-19 mortality: a population-based study of 11.8 million people in Ontario, Canada. Clin Infect Dis. 2023;76(6):1110–1120.

23. Palis H, Bélair MA, Hu K, Tu A, Buxton J, Slaunwhite A. Overdose deaths and the COVID-19 pandemic in British Columbia, Canada. Drug Alcohol Rev. 2022;41(4):912–917.

24. Pezzullo AM, Axfors C, Contopoulos-Ioannidis DG, Apostolatos A, Ioannidis JPA. Age-stratified infection fatality rate of COVID-19 in the non-elderly population. Environ Res. 2023;216(Pt 3):114655.

25. Nepomuceno MR, Klimkin I, Jdanov DA, et al. Sensitivity analysis of excess mortality due to the COVID-19 pandemic. Population and Development Review 2022;48(2):279–302.

26. Stang A, Standl F, Kowall B, et al. Excess mortality due to COVID-19 in Germany. J Infect. 2020;81(5):797–801.

27. Levitt M, Zonta F, Ioannidis JPA. Excess death estimates from multiverse analysis in 2009-2021. Eur J Epidemiol. 2023, https://doi.org/10.1007/s10654-023-00998-2

28. Ioannidis JP, Zonta F, Levitt M. Flaws and uncertainties in pandemic global excess death calculations. Eur J Clin Invest. 2023;53:e14008.

29. Nadarajah R, Wu J, Hurdus B, Asma S, Bhatt DL, Biondi-Zoccai G, Mehta LS, Ram CVS, Ribeiro ALP, Van Spall HGC, Deanfield JE, Lüscher TF, Mamas M, Gale CP. The collateral damage of COVID-19 to cardiovascular services: a meta-analysis. Eur Heart J. 2022;43(33):3164–3178.

30. Khanijahani A, Iezadi S, Gholipour K, Azami-Aghdash S, Naghibi D. A systematic review of racial/ethnic and socioeconomic disparities in COVID-19. Int J Equity Health. 2021;20(1):248.

31. Cajachagua-Torres KN, Quezade-Pinedo HG, Huayaney-Espinoza CA, Obeso-Manrique JA, Pena-Rodriguez VA, Vidal E, Huicho L. COVID-19 and drivers of excess death rate in Peru: a longitudinal ecological study. Heliyon 2022;8:e11948.

32. Axfors C, Ioannidis JPA. Infection fatality rate of COVID-19 in community-dwelling elderly populations. Eur J Epidemiol. 2022;37(3):235–249.

33. Wilmoth JR, Andreev K, Jdanov D, et al. Methods protocol for the human mortality database. University of California, Berkeley, and Max Planck Institute for Demographic Research, Rostock. URL: http://mortality.org [version 31/05/2007], 9, pp.10–11.

34. Scholey J. Robustness and bias of European excess death estimates in 2020 under varying model specifications. 2021, medRxiv: https://www.medrxiv.org/content/10.1101/2021.06.04.21258353v1

35. Islam N, Jdanov DA, Shkolnikov VM, Khunti K, Kawachi I, White M, Lewington S, Lacey B. Effects of Covid-19 pandemic on life expectancy and premature mortality in 2020: time series analysis in 37 countries. BMJ. 2021;375:e066768.

36. Nemeth L, Jdanov DA, Shkolnikov VM. An open-sourced, web-based application to analyze weekly excess mortality based on the Short-term Mortality Fluctuations data series. PLoS One. 2021;16(2):e0246663.

37. Preston SH. The changing relation between mortality and level of economic development. Popul Stud (Camb). 1975;29:231–48.

38. World Bank indicators, in: https://data.worldbank.org/indicator/NY.GDP.PCAP.CD?most_recent_value_desc=true, last accessed March 26, 2023.

39. Income Distribution Database. In: OECD.org, measure: Gini (disposable income, post taxes and transfers), last accessed March 26, 2023.

40. World Bank indicators, in: https://data.worldbank.org/indicator/SI.POV.GINI/)

41. In: https://en.wikipedia.org/wiki/List_of_sovereign_states_by_percentage_of_population_living_in_poverty, last accessed March 26, 2023.

42. Islam N, Jdanov DA. Age and sex adjustments are critical when comparing death rates. BMJ. 2023;381:p845.

